# Midlife mobility assessment from the pocket: agreement and consistency between smartphone placement configurations

**DOI:** 10.64898/2026.11.05.26367337

**Authors:** Dan Keren, Roee Hayek, Itai Gutman, Yaniv Nudelman, Shmuel Springer

## Abstract

Subtle declines in mobility may emerge during midlife and can be detected using smartphone-based motion measures. Instrumented assessment of balance and functional tasks commonly uses lower-back placement, which requires device fixation and may limit convenient real-world monitoring. This study compared mobility outcomes obtained from a practical front-pocket/thigh placement with those from a standardized lower-back placement during static balance, sit-to-stand (STS), and Timed Up and Go (TUG) tests. Forty-six adults aged 40-64 years completed static balance, 30-second STS, and TUG assessments during simultaneous smartphone recording at the lower back and pocket/thigh. Outcomes included two- and three-dimensional sway, STS repetition count, average repetition duration, lower limb power, and maximal vertical velocity, as well as TUG total duration and peak yaw angular velocity during both turns. Associations, intraclass correlation coefficients, and Bland-Altman analyses were used to characterize the relationship and agreement between the two placements. Balance outcomes showed strong correlations between placements (r_s_=0.83-0.93), with moderate-to-excellent consistency for most conditions, although pocket/thigh sway values were systematically higher. TUG total duration showed strong correlation and good absolute agreement (r_s_=0.895; ICC=0.881), and turning velocity showed good consistency during the first and the second turns. STS repetition count and average repetition duration demonstrated strong correlations between placements (r=0.996 and 0.985, respectively), with excellent absolute agreement for repetition count and excellent consistency for average repetition duration (ICC=0.993 and 0.982, respectively). In contrast, STS power and maximal vertical velocity showed weaker correlations and consistency (r=0.709 and r_s_=0.382; ICC=0.600 and 0.417, respectively). Pocket/thigh smartphone placement supports practical mobility assessment across balance, STS and TUG. Although some movement-derived measures showed placement-dependent differences in absolute values, relative performance patterns were generally preserved across placements. These findings support the development of pocket-based smartphone assessment for simplified and potentially unsupervised mobility monitoring in midlife, while placement-sensitive outcomes require further validation against external reference measures.

**Author Summary:** Wearable technologies, including smartphones, offer promising ways to assess mobility during midlife, when subtle changes in physical performance may begin to emerge but go undetected by conventional clinical tests. However, most research using these technologies has been conducted under controlled conditions, with devices often attached to the lower back. Although this approach provides useful measurements, it requires fixation, making it less practical for everyday use.

In this study, we examined whether smartphones carried in a front pocket could provide mobility measurements comparable to those obtained from the lower back. Forty-six middle-aged adults completed balance, chair-standing, and walking and turning tasks while smartphones recorded their movements at both locations.

We found that measures of mobility, such as chair-standing repetitions and walking task completion time, were highly comparable between smartphone locations. Balance measurements, such as body sway while standing on one leg, showed similar patterns, although their absolute values differed. In contrast, measures of muscle power and maximum upward movement speed during chair-standing were more sensitive to smartphone placement.

Our findings support the potential of pocket-based smartphone assessment as a practical approach to measuring mobility beginning in midlife, while further research is needed to establish its suitability for everyday use.

## Background

Midlife represents a critical stage in the aging trajectory, during which subtle and often undetected declines in mobility may begin to emerge, even when overall health is largely preserved [1]. Changes in key mobility-related domains, including gait, balance, and lower-limb strength and power, may represent early manifestations of functional decline [1–5]. Accordingly, there is a growing need for accessible, sensitive, and convenient assessment methods capable of detecting early declines in locomotion performance during midlife.

Traditional assessments, typically based on simple clinical outcomes such as walking speed or sit-to-stand (STS) duration, may lack the sensitivity required to detect early mobility deterioration in midlife [2–4]. Recent studies have shown that instrumented assessments using wearable inertial sensors can overcome some of the ceiling limitations of conventional measures and detect subtle differences among community-dwelling middle-aged adults [2–4,6]. These sensor-derived measures may provide added sensitivity by capturing specific components of postural control and movement quality that are not reflected in conventional outcomes. For example, smartphone-derived balance measures based on postural sway differentiated middle-aged from younger adults [2]; Timed-Up-and-Go (TUG) derived turning velocity was associated with early functional decline despite limited sensitivity of total TUG duration [6]; and smartphone-derived STS power distinguished middle-aged from younger adults when conventional STS duration did not [4].

Although wearable technologies, including smartphones, have shown promising results, the existing evidence is largely based on studies conducted under controlled research conditions, using standardized sensor configurations typically placed at the lower back to ensure signal consistency [3,4,7–12]. While this placement provides a stable representation of whole-body movement, it requires dedicated fixation during measurement, which may limit real-world implementation and ecological validity. A front-pocket configuration better reflects everyday smartphone carrying habits and may facilitate independent use by eliminating the need for dedicated mounting accessories or assistance with device placement, thereby supporting future unsupervised assessment. However, because the pocket-mounted device captures motion farther from the body’s center of mass and from a different body segment, sensor-derived outcomes may differ from those obtained at the lower back [13], highlighting the need to evaluate the relationship between measurements obtained from the two configurations. Existing validation studies that examined smartphone-based mobility assessment have primarily involved younger or older adults [7,13,14], with limited investigation in middle-aged populations, despite the variability in functional performance characteristics in this age group [1,2]. Moreover, these studies have typically assessed single mobility domains rather than a comprehensive set of outcomes [8,11,13,14].

OneStep is a smartphone-based platform that uses embedded inertial sensors and machine-learning algorithms to assess mobility without requiring dedicated external sensors [14–17]. Previous validation studies have demonstrated its ability to measure spatiotemporal gait parameters using a smartphone carried in the pocket, with generally good agreement with reference gait-analysis systems [14,15]. Additional studies have supported its application for gait assessment under clinical and real-world recording conditions [16,17]. However, evidence regarding pocket-based assessment has focused primarily on gait, with limited investigation in middle-aged adults and little evidence regarding other mobility domains, including balance, STS, and TUG, which may help detect early mobility deterioration in midlife.

This study aimed to compare mobility metrics obtained from practical pocket/thigh and standardized lower-back smartphone placements and to quantify placement-related differences across static balance, TUG, and STS tasks in community-dwelling middle-aged adults. We hypothesized that pocket/thigh-derived measurements would show moderate-to-strong associations and good-to-excellent consistency with lower-back-derived measurements. For conventional clinical outcomes, specifically STS repetition count and TUG total duration, we expected good-to-excellent absolute agreement between placements, as these outcomes are less dependent on sensor location.

## Methods

### Participants

An a priori sample size calculation for the planned ICC analysis indicated that 45 participants were required, assuming a minimum acceptable ICC of 0.50, an expected ICC of 0.75, two measurements per participant, α = 0.05, and 80% power [18]. Accordingly, 46 community-dwelling middle-aged adults were recruited.

Participants were included if they were 40-64 years old, lived independently in the community, and were able to walk outdoors without assistance. Exclusion criteria included neurological, orthopedic, vestibular, or visual conditions that could affect mobility (e.g., stroke, Parkinson’s disease, recent fracture or orthopedic surgery, age-related macular degeneration, glaucoma, or diabetic retinopathy).

The study was approved by the Ariel University Ethics Committee (AU-HEA-SS−20230806), All research was conducted in accordance with the pre-approved study protocol, applicable institutional guidelines and regulations, and the ethical principles of the Declaration of Helsinki. All participants provided written informed consent prior to participation.

### Experimental Setup

Data were collected simultaneously from two primary smartphone placements, a standardized lower-back placement [3,4,13], with the smartphone secured in a dedicated pouch over the lumbar region, and a dominant-side front-pocket/thigh placement, with the smartphone carried in the front pants pocket when available, or in a snug pouch over the anterolateral thigh on the front-pocket region, when a suitable pocket was unavailable. For the TUG task only, an additional smartphone was placed at the non-dominant front-pocket/thigh location so that data from the pivot-leg side could be analyzed regardless of the participant’s self-selected turning direction, as schematically illustrated in Fig 1.

**Fig 1.**
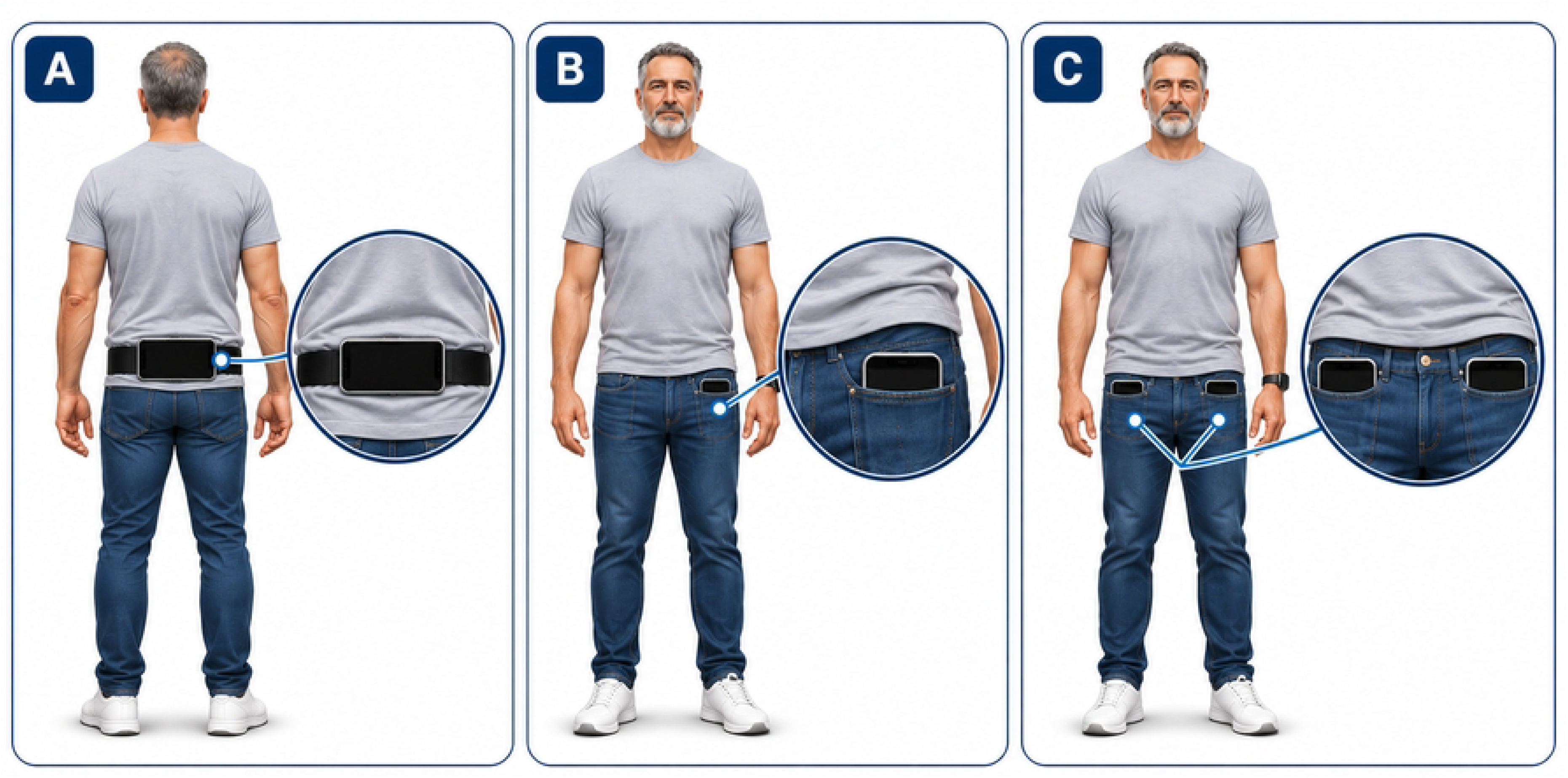
Smartphone placement configurations used during mobility assessments. (A) Horizontal smartphone placement over the lumbar region using a dedicated pouch. (B) Dominant-side front-pocket/thigh placement. (C) Bilateral front-pocket/thigh placement used during the Timed Up and Go test to enable analysis of turning-related measures from the pivot-leg side.

To reduce potential device-specific bias, smartphone device assignment to body location was randomized across participants. The devices used were two Xiaomi Mi A3 smartphones (Android 11; approximate dimensions 153.5 × 71.9 × 8.5 mm; weight 173 g) and one Samsung Galaxy S23 smartphone (Android 15; approximate dimensions 146.3 × 70.9 × 7.6 mm; weight 168 g).

All smartphones recorded inertial data simultaneously using OneStep’s smartphone gait and mobility analysis system (software versions 4.0.#; Celloscope Ltd., Tel Aviv, Israel), which captured tri-axial linear acceleration and angular velocity from the embedded accelerometer and gyroscope, along with device timestamps [14]. Recordings from the relevant device locations were uploaded to the OneStep platform and processed using a standardized analysis pipeline. Before each trial, participants performed three steps in place, generating a predefined acceleration marker that was used to temporally align recordings from the two smartphone placements using signal-correlation-based matching. Participants were identified in the OneStep application and platform only by their unique study identification number.

### Procedure and Outcome Measures

All assessments were conducted in a single supervised session following a standardized protocol. Tasks were performed in the following order: static balance, TUG, and 30-second STS. The STS was performed last to minimize the potential influence of fatigue on the preceding assessments. Task initiation and termination were guided by standardized audio cues (“GO” and “recording stopped”).

In cases of technical recording issues (e.g., incorrect recording initiation or inadequate device fixation), or incorrect task execution due to misunderstanding of the instructions or delayed response to the start cue, participants were permitted a second attempt. For static balance tasks, a second attempt was also permitted when the task was not completed because of loss of balance. If neither attempt was completed successfully, the better performance was retained for analysis.

Balance was assessed under four standing conditions: (a) tandem stance with eyes open, performed for 30 seconds with the dominant foot behind the non-dominant foot and hands on hips; (b) tandem stance with eyes closed, performed in the same position for 10 seconds; (c) single-leg stance, performed for 30 seconds on the dominant leg with hands on hips and without contact between the legs; and (d) standing on a foam surface with eyes closed for 30 seconds in a narrow stance. For each balance condition, two-dimensional (2D) sway dispersion, quantified using the 95% confidence ellipse (m/s²)², and three-dimensional (3D) sway dispersion, quantified using the 95% confidence ellipsoid (m/s^2^)^3^ were extracted as outcome measures.

For the TUG assessment, participants began seated on a standard chair, stood without using their hands, walked three meters at their fastest safe speed without running, turned around a predefined marker, returned to the chair, and sat down. TUG outcomes included total task duration (s), and peak yaw angular velocity during the first and second turns (°/s) [6,19].

In the 30-second STS task, participants began seated on a standard chair and completed as many repeated sit-to-stand cycles as possible. Arms were crossed over the shoulders throughout the test. Each repetition required full knee and hip extension during standing and complete contact with the chair while sitting, ensuring standardized movement execution. STS outcome measures included repetition count (n), average repetition duration (s), average lower-limb power (W), and maximal vertical velocity during the rising phase (m/s).

### Signal Processing

Inertial signals were sampled at 100 Hz and processed using the standardized OneStep analysis pipeline. Signals from both smartphone placements were processed using the same general analytical approach, with placement-specific adjustments made where required to account for differences in signal characteristics. Linear acceleration signals, with the gravitational component removed by the smartphone’s sensor-fusion algorithm, were used for subsequent processing. For balance assessments, 2D and 3D sway dispersion were calculated from the covariance matrices of the mediolateral and anteroposterior acceleration components and the full three-dimensional acceleration signal, respectively [2]. Eigenvalues of the respective covariance matrices were scaled according to the 95% χ² confidence region and used to calculate the area of the 95% confidence ellipse and volume of the 95% confidence ellipsoid.

For TUG, pocket/thigh sub-phases were identified from yaw and pitch signals using windowed angular-displacement thresholds, with minimum cumulative angular changes of 105° for turns and 60° for sit-to-stand and stand-to-sit transitions. Turn boundaries were refined using a fourth-order zero-phase Butterworth low-pass filter with a 1.5-Hz cutoff. Peak yaw angular velocity was derived using a Savitzky-Golay filter (101-sample window, fourth-order polynomial, first derivative) and calculated within each segmented turn. For lower-back recordings, the previously validated TUG algorithm was applied [6], with turns identified from filtered yaw angular velocity and postural transitions from pitch angular velocity. Total TUG duration was calculated from the onset of standing to completion of the final sitting phase.

For STS, repetitions were identified from cyclic changes in the smoothed pitch-angle signal using prominence-based peak detection. For pocket/thigh recordings, gravity-free acceleration was rotated to the earth frame to obtain vertical acceleration, detrended, and numerically integrated to derive vertical velocity. Rising and sitting phases were identified from positive and negative vertical velocities, respectively. Maximal vertical velocity was defined as the peak velocity during the rising phase, and lower-limb power was calculated as the product of body mass, vertical acceleration including gravity, and vertical velocity, averaged across repetitions. For lower-back recordings, the previously established processing approach was applied [4], with vertical acceleration and pitch angular velocity filtered using a fourth-order Butterworth filter with a 1.5-Hz cutoff. Vertical acceleration was integrated with polynomial detrending to obtain vertical velocity and rising and sitting phases were identified from the corresponding velocity peaks.

Recordings with incomplete data, unclear synchronization markers, non-informative signals, or unreliable task segmentation were excluded from the relevant task-specific analysis.

### Statistical Analysis

Descriptive statistics were reported as mean ± standard deviation (SD) and median with interquartile range (IQR) for demographic and outcome measures. Distributional characteristics of continuous variables were assessed by visual inspection of histograms and Q-Q plots.

Associations between lower-back and pocket/thigh measurements were assessed separately for each task-specific outcome using Pearson or Spearman correlation coefficients, according to the distributional characteristics of each outcome. Correlation coefficients were interpreted using commonly applied thresholds, with values <0.30 considered weak, 0.30-0.50 moderate, and >0.50 strong [20]. The lower-back placement was compared with the dominant-side pocket/thigh placement for balance and STS outcomes and with the pivot-leg pocket/thigh placement for TUG outcomes.

Agreement and consistency between placements were quantified using single-measure intraclass correlation coefficients (ICC) based on a two-way mixed-effects model. Absolute agreement ICCs were used for TUG total duration and STS repetition count because these outcomes were expected to yield equivalent values across placements. Consistency ICCs were used for each signal-derived outcome to assess whether relative between-participant differences were preserved across placements despite potential differences in absolute magnitude. ICC values were interpreted as poor (<0.50), moderate (0.50-0.75), good (0.75-0.90), and excellent (>0.90) [21,22].

Bland-Altman analyses were used to quantify systematic bias and variability between placements. Bias was calculated as the lower-back measurement minus the pocket/thigh measurement; therefore, positive values indicate higher measurements at the lower-back placement, whereas negative values indicate higher measurements at the pocket/thigh placement. The limits of agreement (LoA) were calculated as the mean bias ± 1.96 × the standard deviation of the differences.

Outcome-specific analyses included all participants with valid measurements. Due to technical recording failures, one participant had missing data for all balance outcomes and STS power, and a different participant had missing pocket/thigh data for all TUG outcomes. Accordingly, analyses included 45 participants for all balance outcomes, STS power, and TUG outcomes, and 46 participants for the remaining STS outcomes.

All statistical analyses were conducted in R (version 4.3.3), with a significance level of α = 0.05. For correlation analyses, Bonferroni correction was applied separately within each task category to account for multiple outcome comparisons. Accordingly, Bonferroni-corrected significance thresholds were α = 0.006 for balance outcomes, α = 0.017 for TUG outcomes, and α = 0.013 for STS outcomes.

## Results

Participant characteristics and descriptive outcome measures for the lower-back and pocket/thigh smartphone placements are presented in Table 1. The final sample included 46 participants (67.4% women), with a mean age of 51.15 ± 6.97 years and mean BMI of 25.69 ± 3.52 kg/m².

**Table 1.** Participant characteristics and descriptive statistics for outcome measures by smartphone placement.

| **Characteristics** |  | **Mean ± SD / Percentage** |  |
|---|---|---|---|
| **Age** (years) |  | 51.15 ± 6.97 |  |
| **Sex,** n (%) |  | Female: 31 (67.4) \| Male: 15 (32.6) |  |
| **Height** (m) |  | 1.67 ± 0.09 |  |
| **Weight** (kg) |  | 71.99 ± 13.55 |  |
| **BMI** (kg/m<sup>2</sup>) |  | 25.69 ± 3.52 |  |
| **Measure** | **outcome** | **Lower-back** | **Pocket/thigh** |
| **Tandem stance, eyes closed** | 2D ellipse (m/s²)² | 25.46 ± 126.06 | 34.69 ± 116.34 |
|  | 3D ellipsoid (m/s²)³ | 32.78 ± 200.73 | 71.40 ± 410.48 |
| **Tandem stance, eyes open** | 2D ellipse (m/s²)² | 0.79 ± 0.92 | 2.27 ± 3.31 |
|  | 3D ellipsoid (m/s²)³ | 0.09 ± 0.17 | 0.27 ± 0.61 |
| **Soft surface, eyes closed** | 2D ellipse (m/s²)² | 0.54 ± 0.60 | 0.66 ± 0.69 |
|  | 3D ellipsoid (m/s²)³ | 0.04 ± 0.06 | 0.07 ± 0.13 |
| **Single-leg stance** | 2D ellipse (m/s²)² | 16.17 ± 55.53 | 31.92 ± 82.47 |
|  | 3D ellipsoid (m/s²)³ | 15.38 ± 74.62 | 34.76 ± 145.62 |
| **TUG** | Total duration (sec) | 6.84 ± 0.97 | 6.46 ± 0.94 |
|  | Peak yaw turn 1 (°/s) | 194.89 ± 34.00 | 203.25 ± 32.04 |
|  | Peak yaw turn 2 (°/s) | 265.38 ± 48.17 | 279.57 ± 47.54 |
| **30-s STS** | Repetitions (n) | 21.09 ± 5.65 | 21.48 ± 5.85 |
|  | Average duration of one repetition (s) | 1.41 ± 0.38 | 1.45 ± 0.41 |
|  | Power (W) | 397.11 ± 171.77 | 216.53 ± 91.88 |
|  | Maximal vertical velocity during standing (m/s) | 0.79 ± 0.19 | 0.48 ± 0.22 |

Table 1 represents summary of participant characteristics and outcome measures. Values are presented as mean ± standard deviation (SD) or percentage, as appropriate. 2D ellipse and 3D ellipsoid represent postural sway measures obtained during standing balance tasks. STS: sit-to-stand; TUG: Timed Up and Go; N: number; s: seconds; W: watts; m: meters; °: degrees.

### Static balance Outcomes

The static balance outcomes are presented in Table 2. Spearman’s rank correlations between lower-back and pocket/thigh placements were strong across all balance conditions and for both 2D ellipse and 3D ellipsoid outcomes (r_s_ = 0.825-0.933, all p < 0.001).

**Table 2.** Correlation, ICC, and Bland–Altman estimates between lower-back and pocket/thigh smartphone placements.

| **Task** | **Outcome** | **N** | **Correlation** | **P** | **ICC** | **ICC 95% CI** | **Bias** | **LoA** |
|---|---|---|---|---|---|---|---|---|
| **Tandem stance, eyes closed** | 2D ellipse (m/s²)² | 45 | rs = 0.866 | <.001 | 0.966** | 0.940-0.982 | -9.23 | −70.64 to 52.18 |
|  | 3D ellipsoid (m/s²)³ | 45 | rs = 0.933 | <.001 | 0.787** | 0.643–0.877 | -38.62 | −452.20 to 374.96 |
| **Tandem stance, eyes open** | 2D ellipse (m/s²)² | 45 | rs = 0.825 | <.001 | 0.466** | 0.203–0.667 | -1.48 | −6.40 to 3.45 |
|  | 3D ellipsoid (m/s²)³ | 45 | rs = 0.899 | <.001 | 0.477** | 0.216–0.674 | -0.19 | −1.08 to 0.71 |
| **Soft surface, eyes closed** | 2D ellipse (m/s²)² | 45 | rs = 0.842 | <.001 | 0.804** | 0.671–0.888 | -0.11 | −0.90 to 0.68 |
|  | 3D ellipsoid (m/s²)³ | 45 | rs = 0.922 | <.001 | 0.598** | 0.371–0.757 | -0.03 | −0.21 to 0.15 |
| **Single-leg stance** | 2D ellipse (m/s²)² | 45 | rs = 0.833 | <.001 | 0.880** | 0.791–0.932 | -15.75 | −83.34 to 51.83 |
|  | 3D ellipsoid (m/s²)³ | 45 | rs = 0.881 | <.001 | 0.809** | 0.677–0.890 | -19.38 | −159.61 to 120.85 |
| **TUG** | Total duration (s) | 45 | r<sub>s</sub> = 0.895 | <.001 | 0.881<sup>†</sup>* | 0.110–0.966 | 0.40 | −0.16 to 0.95 |
|  | Peak yaw turn 1 (°/s) | 45 | r<sub>s</sub> = 0.790 | <.001 | 0.822** | 0.697–0.898 | -8.07 | −46.95 to 30.80 |
|  | Peak yaw turn 2 (°/s) | 45 | r<sub>s</sub> = 0.849 | <.001 | 0.837** | 0.723–0.907 | -14.76 | −68.46 to 38.94 |
| **30-s STS** | Repetitions (n) | 46 | r = 0.996 | <.001 | 0.993<sup>†</sup>** | 0.975–0.997 | -0.39 | −1.52 to 0.74 |
|  | Average duration of one repetition (s) | 46 | r = 0.985 | <.001 | 0.982** | 0.967-0.990 | -0.05 | -0.19 to 0.10 |
|  | Power (W) | 45 | r = 0.709 | <.001 | 0.600** | 0.375–0.758 | 178.48 | −64.46 to 421.43 |
|  | Maximal vertical velocity during standing (m/s) | 46 | r<sub>s</sub> = 0.382 | .009 | 0.417* | 0.147–0.629 | 0.31 | −0.11 to 0.74 |

Table 2 summarizes paired sample sizes, correlation coefficients, ICC estimates, mean bias, and limits of agreement for all task-specific outcomes. Values are based on paired observations for each outcome. r<sub>s</sub> denotes Spearman’s rank correlation coefficient, and r denotes Pearson’s correlation coefficient. All reported correlations remained statistically significant after Bonferroni correction. ICC values are single-measure estimates from a two-way mixed-effects model consistency ICCs were used unless otherwise indicated. <sup>†</sup> Indicates absolute agreement ICCs. For ICC estimates, \* p < 0.05 and \*\*p < 0.001. Bias was calculated as lower-back minus pocket/thigh values, with positive values indicating higher lower-back estimates and negative values indicating higher pocket/thigh estimates. p values are unadjusted. LoA, limits of agreement; TUG, Timed Up and Go; STS, sit-to-stand; n: number; s: seconds; W: watts; m: meters; °: degrees.

Consistency ranged from poor to excellent across the static balance outcomes (ICC = 0.466-0.966). The lowest consistency was observed during tandem stance with eyes open (ICC = 0.466 and 0.477 for the 2D ellipse and 3D ellipsoid outcomes, respectively), whereas the remaining outcomes showed moderate-to-excellent consistency (Fig 2). Bland-Altman analyses showed consistent negative bias across all static balance outcomes, indicating higher sway-dispersion values at the pocket/thigh placement than at the lower-back placement. Bias and 95% limits of agreement are also presented in Table 2.

**Fig 2.**
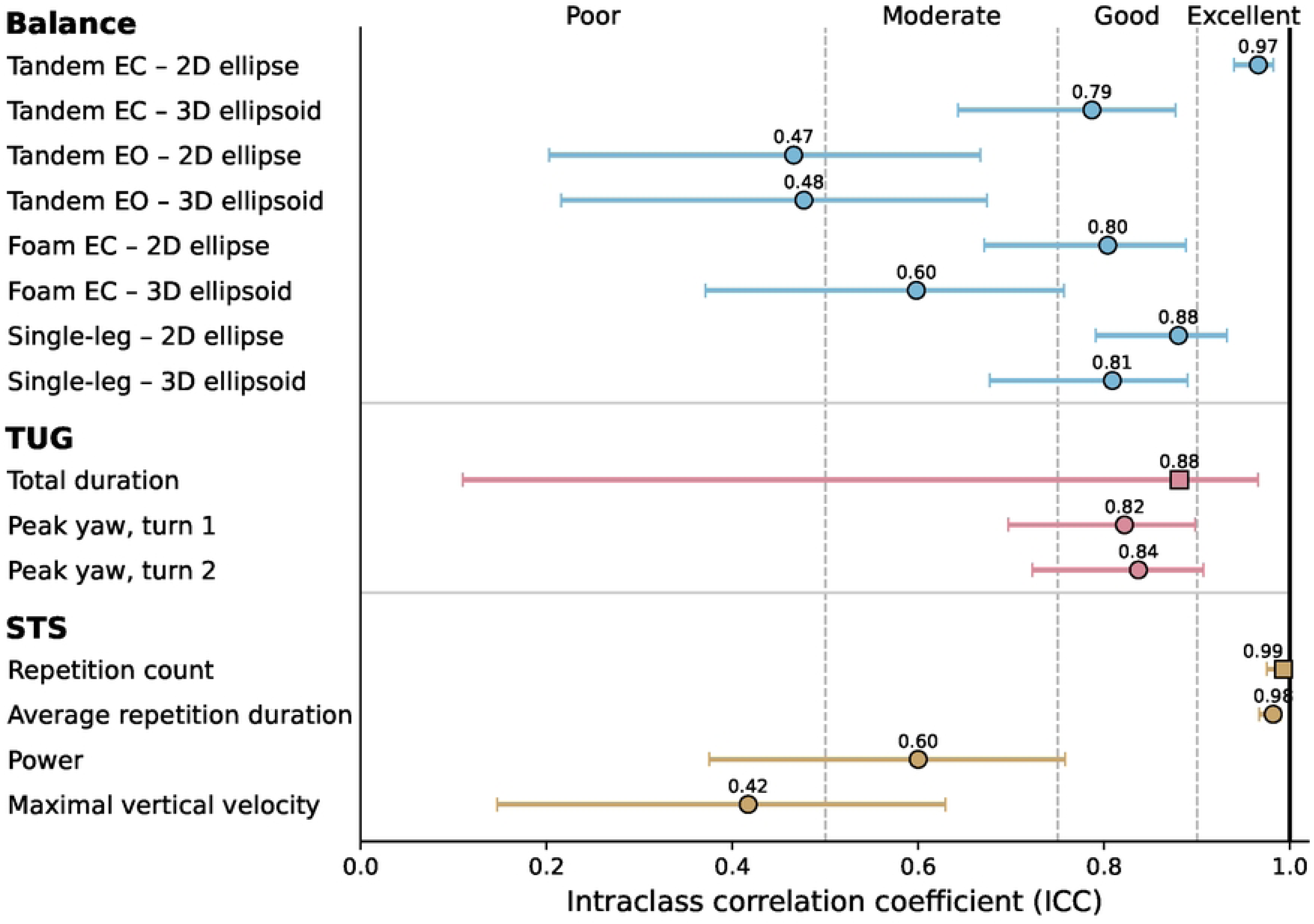
Agreement and consistency between smartphone placements. Intraclass correlation coefficient (ICC) estimates with 95% confidence intervals for agreement or consistency between lower-back and pocket/thigh smartphone placements across balance, Timed Up and Go (TUG), and sit-to-stand (STS) outcomes. Circle markers indicate consistency ICCs, whereas square markers indicate absolute-agreement ICCs. Vertical reference lines denote conventional interpretation thresholds for ICC magnitude: poor (<0.50), moderate (0.50–0.75), good (0.75–0.90), and excellent (>0.90). Values above each marker indicate the ICC point estimate. EC: eyes closed; EO: eyes open.

### TUG Outcomes

TUG total duration demonstrated strong correlation between lower-back and pocket/thigh placements (rs = 0.895), and good absolute agreement (ICC = 0.881), although the 95% CI was wide (0.110-0.966). Peak yaw angular velocity demonstrated strong correlation during turn 1 (r_s_ = 0.790) and turn 2 (r_s_ = 0.849). Consistency was good for turn 1 (ICC = 0.822, 95% CI 0.697-0.898), and for turn 2 (ICC = 0.837, 95% CI 0.723-0.907) (Fig 2). Bland-Altman analyses indicated a small positive bias for TUG total duration, whereas peak yaw angular velocity showed negative biases and relatively wide limits of agreement during both turns. Complete correlation, ICC, bias, and limits of agreement results are presented in Table 2.

### STS Outcomes

STS repetition count and average repetition duration demonstrated strong correlations between lower-back and pocket/thigh placements (r = 0.996 and 0.985, respectively).

Repetition count showed excellent absolute agreement (ICC = 0.993), while average repetition duration showed excellent consistency (ICC = 0.982). STS power showed strong correlation and moderate consistency between placements (r = 0.709; ICC = 0.600), with the 95% ICC confidence interval ranging from poor to good. Maximal vertical velocity showed moderate correlation and poor consistency (r = 0.382; ICC = 0.417), with the 95% ICC confidence interval ranging from poor to moderate (Fig 2). Bland-Altman analyses indicated minimal bias for repetition count and average repetition duration, whereas greater systematic differences were observed for power and maximal vertical velocity. Complete correlation, ICC, bias, and 95% limits of agreement results are presented in Table 2.

## Discussion

The present study examined whether mobility outcomes derived from a practical front-pocket/thigh smartphone placement showed comparable measurement patterns to those obtained from a standardized lower-back placement in community-dwelling middle-aged adults. As hypothesized, all static balance outcomes showed strong associations between the two smartphone placements, although consistency varied across conditions and was lower for tandem stance with eyes open. STS repetition count and TUG total duration also supported our hypothesis for conventional clinical outcomes, demonstrating excellent and good absolute agreement between placements, respectively. In contrast, consistency between placements varied across the other outcomes. STS power and maximal vertical velocity showed lower consistency between placements, whereas turning velocity during the TUG showed good consistency across the two turns. These findings indicate that values obtained from the two placements may not be interchangeable for all outcomes.

For static balance, participants who showed greater sway at the lower-back placement generally also showed greater sway at the pocket/thigh placement, despite differences in absolute sway values. However, consistency varied across conditions, and it was lower during tandem stance with eyes open. The availability of visual input may have reduced the postural challenge [23], and consequently, the variability in performance between participants. Because ICC values are influenced by between-participant variability [24], this may help explain the lower consistency observed in this condition. The pocket/thigh recordings also produced higher sway values. This may reflect greater movement of the thigh relative to the lower back during postural adjustments, resulting in larger accelerations recorded at the thigh. Sensor location along a body segment has previously been shown to influence acceleration magnitude, supporting the possibility that the higher sway values reflect the biomechanical characteristics of the measurement location rather than poorer measurement performance [25]. Thus, pocket/thigh placement may be useful for practical balance assessment and monitoring, but absolute sway values obtained from the two placements should not be used interchangeably.

TUG outcomes showed good agreement and consistency between smartphone placements. Total duration demonstrated good absolute agreement, while turning velocity showed good consistency across both turns, indicating that the practical pocket/thigh placement captured TUG performance in a manner broadly comparable with the lower-back placement. The good agreement observed for total duration was consistent with our hypothesis, as this outcome reflects completion time of the entire task rather than the magnitude of segment-specific angular motion. The wide confidence interval for TUG total duration may reflect limited between-participant variability, which can affect ICC precision [26], as well as small placement-related differences in detecting task completion due to variations in turn-to-sit strategy and sensor location [27,28]. Turning velocity demonstrated good consistency across both turns, despite the potential influence of sensor placement and variation in segmental motion during turning. Previous research has shown that sensor-derived measurements can be influenced by sensor placement and that this effect may depend on the specific functional movement being analyzed [29]. In the present study, bilateral pocket/thigh recordings allowed turning velocity to be derived from the pivot-leg side, based on evidence that the inner and outer limbs have different biomechanical roles during turning [30,31]. However, practical assessment with a single smartphone would require consideration of both carrying side and turning direction. Future studies should determine whether pocket-derived turning measures remain valid when the pivot-leg side cannot be identified.

The STS findings further illustrate that the influence of smartphone placement depends on the specific outcome being measured. Repetition count and average repetition duration were highly consistent between placements, whereas power and maximal vertical velocity showed substantially weaker consistency. This difference likely reflects the nature of these measures. Repetition count and duration primarily capture the timing and completion of the movement, whereas power and maximal vertical velocity are more directly influenced by the movement of the body segment on which the smartphone is located. During the rising phase, the lower-back smartphone primarily captures trunk motion closer to the body’s center of mass, whereas the pocket/thigh smartphone captures thigh motion and hip flexion-extension dynamics. Consequently, power and maximal vertical velocity derived from the two placements may represent different aspects of the STS movement and should not be used interchangeably.

Beyond the comparison between smartphone placements, an important finding of the present study is the feasibility of obtaining smartphone-derived measures across multiple mobility domains using a practical pocket/thigh configuration in community-dwelling middle-aged adults. This extends the potential role of smartphone-based assessment from controlled testing toward more accessible monitoring of mobility beginning in midlife. The good agreement and consistency observed between smartphone placements across several balance, STS, and TUG outcomes support the potential use of practical front-pocket placement for mobility assessment without dedicated mounting accessories. However, this potential appears to be outcome-specific, as STS power and maximal vertical velocity were more sensitive to measurement location and should not be used interchangeably across placements. Given that mobility measures in midlife have been associated with future disability [32,33], the ability to obtain instrumented mobility measures from a smartphone carried in the pocket may provide an important basis for developing accessible approaches to earlier mobility monitoring. Future studies should examine its application in field-based and unsupervised settings, as the ecological validity of pocket-based smartphone mobility assessment outside supervised testing remains to be established.

The results of this study should be interpreted in light of several limitations. First, the pocket/thigh configuration was not completely uniform. Participants used their own front pockets when suitable, whereas a snug pouch positioned over the front-pocket region of the thigh was used when a suitable pocket was unavailable. This pragmatic approach introduced variability in device positioning and pocket characteristics, but also better reflected the intended practical use of the assessment. Outcomes that remained highly consistent across placements despite this variability may therefore be more robust to differences in everyday smartphone carrying conditions. Similarly, the use of different smartphone models, with devices randomized across measurement locations, was intended to reduce dependence on a specific device configuration and increase the practical relevance of the findings. Second, all assessments were conducted during a single supervised session. Thus, although the findings support the feasibility of pocket-based smartphone assessment, they do not establish its performance during independent or repeated assessment in real-world settings. Finally, the relatively modest sample consisted of community-dwelling middle-aged adults able to walk independently, which may limit generalizability to individuals with greater functional limitations.

## Conclusions

This study demonstrates the feasibility of obtaining smartphone-derived measures across multiple mobility domains using a practical pocket/thigh configuration in community-dwelling middle-aged adults. Pocket/thigh measurements demonstrated good agreement or consistency with lower-back measurements for most balance, STS, and TUG outcomes.

These findings support the further development of pocket-based smartphone assessment as an accessible approach for mobility assessment and monitoring during midlife, with future studies needed to establish its performance during independent and real-world use.

## Data Availability

De-identified participant-level data underlying the findings of this study may be made available upon reasonable request, subject to approval by the Ariel University Ethics Committee and in accordance with participant consent and applicable privacy regulations. Requests may be initiated through the corresponding author, with data access subject to institutional review and approval.

## Notes

### Competing Interest Statement

The authors have declared no competing interest.

### Author Declarations

The study was approved by the Ariel University Ethics Committee (Approval No. AU-HEA-SS-20230806). All participants provided written informed consent prior to participation. The study was conducted in accordance with the Declaration of Helsinki.

